# Assessing the performance of Laboratory professionals following a day of malaria microscopy training in Kano, Nigeria

**DOI:** 10.1101/2024.02.23.24303252

**Authors:** Oluwaseunayo Deborah Ayando, Nirmal Ravi, Okoronkwo Marizu Obinnaya

## Abstract

Malaria is a global health burden with a projection of 247 million cases in 2021 in 84 countries known to be malaria-endemic. The majority of the cases are expected to occur in WHO African Region countries inclusive of Nigeria where the disease is a public health concern particularly in the northern regions. This study aimed to evaluate the effectiveness of a one-day malaria microscopy training program for laboratory professionals in Kano, Nigeria, and to assess the impact of the training on their proficiency in detecting malaria parasites. A total of 56 medical laboratory professionals from both public and private healthcare facilities participated in the training, which was based on the WHO basic microscopy learners guide. The training included theoretical and practical components, focusing on blood film preparation, staining techniques, and identification of Plasmodium species. Participants’ knowledge was assessed before and after the training using a pre-test and post-test survey, and their proficiency in malaria diagnosis was evaluated through the examination of a panel of stained blood slides. The sensitivity and specificity of the participants in detecting malaria parasites were determined as 66% and 34% respectively, indicating moderate sensitivity but very low specificity. A significant improvement in participants’ knowledge of malaria detection was observed post-training, with private facilities showing a median knowledge score increase from 47.5% to 65.0%, and public facilities from 55.0% to 70.0%. However, the overall agreement between participant readers and an expert reader, measured by the kappa coefficient, was zero, suggesting no agreement beyond chance. The study highlighted the need for enhanced training and standardization in malaria microscopy to improve diagnostic accuracy. Despite the moderate increase in knowledge and sensitivity post-training, the low specificity and agreement underscore the importance of ongoing education and quality assurance measures in malaria diagnosis, especially in high-prevalence settings like Kano.

## Introduction

Malaria is an infectious, vector-transmitted parasitic disease caused by the protozoan *Plasmodium* species. *P.falciparum and P.vivax* are the most prevalently infective species compared to *P.malariae, P.ovale, and P.knowlesi* [1]. Parasites are transmitted as a sporozoites to humans through a blood meal by a female *Anopheles* mosquito. Through a complex human-mosquito life cycle [2], *Plasmodium* species infect hepatocytes and then erythrocytes, causing nonspecific acute febrile symptoms [3]. Symptoms arise 10-15 days post-infection and can lead to severe illness or death if treatment is not administered 24-48 hours after symptom onset [4]. Prophylactic and therapeutic medications exist, but no reliable vaccine is available. Recently, the RTS, S/AS01 vaccine that offers partial protection to children [5] was developed and introduced in Malawi [6].

Malaria is a global health burden with a projection of 247 million cases in 2021 in 84 countries known to be malaria-endemic. The majority of the cases are expected to occur in WHO African Region countries inclusive of Nigeria where the disease is a public health concern. The nation accounted for roughly 27% of the world’s malaria cases in 2021, according to the 2021 World Malaria Report, and 31% of all malaria deaths worldwide. It also had the largest number of malaria deaths. All year long, there is a risk of transmission across the nation. However, the northern and northeastern regions of the nation have the greatest rates of malaria [7].

Nigeria, the most populated (203.4 million people) and one of the fastest-growing nations in Africa (2.54% growth rate) [8], has the largest global disease burden of malaria, accounting for approximately a quarter of all persons at risk in 2017 (190.9 million). The Plasmodium falciparum malaria species is the most common in Nigeria. Approximately 67.1% of all Anopheles gambiae s.s. collected nationwide are primary vectors, with Anopheles funestus serving as a secondary vector in certain regions of Nigeria [9]. The intricate interaction between the trinity of mosquitoes, parasites, and humans sustains malaria sickness. The disease’s prevalence could be influenced by human behavior or inactivity. Men have a greater parasite biomass (99%), whereas mosquitoes only have one parasite biomass (1%). In most cases, a greater proportion of the human population in malaria-endemic regions is considered to be “infected” yet asymptomatic. The unintentional parasite reservoirs that sustain the infection’s spread are these asymptomatic groups [10].

Early diagnosis and treatment are the foundation for managing and the key to lowering mortality and morbidity from malaria. Reducing drug waste and potential resistance to antimalarial medications, detecting additional lethal non-malarial sources of illness, and facilitating the efficient treatment of febrile patients are all made possible by accurate diagnosis [11–12]. RDTs are a rather quick and easy method of diagnosing malaria; they are typically utilized in places without electricity and with less competent workers, albeit their cost is a drawback [13]. According to Boadu et al. (2016), patients in urban locations with limited facilities perceive laboratory diagnosis as a waste of time due to lengthy waiting times caused by high caseloads [14]. Many times, testing was done arbitrarily, or for patients who were hospitalized or jailed, therapy was given before the results were known [15].

Microscopy continues to be the gold standard for diagnosing malaria in Nigeria despite these drawbacks. Using light microscopy, Plasmodium parasites can be found in stained blood smears. For the easy and affordable detection and identification of several Plasmodium species stages in peripheral blood smears, microscopy is used. However, this approach takes time and expertise, particularly when dealing with individuals who have low-level parasitemia [16]. Microscopists in many developing nations face enormous workloads, inadequate supervision, and inadequate [17]. Kano State is in the Northern region of Nigeria. The Nigerian Census Bureau projected Kano’s population to be 13 million by 2016 based on 2006 census data [18]. The variation in malaria prevalence detected by RDT (Rapid Diagnostic Tests) and microscopy across all states, with RDT and microscopy results diverging from their weighted averages, suggests RDTs may have a higher incidence of false positives than microscopy (National Malaria Elimination Programme, 2015; Ajumobi et al., 2015; Wongsrichanalai et al., 2019). [18-20). This study therefore sought to assess laboratory professionals’ knowledge and microscopy proficiency in detecting malaria parasites after a microscopy training was conducted.

## Methods

### Study Area

Kano is the municipal center of Kano state in Northcentral Nigeria. The municipality’s total area is 193 square miles and it has the second-largest population in Nigeria following Lagos. The urban population in 2018 was 3.8 million with an estimated growth rate of 3.1% from 2018 to 2030 [21]. Malaria transmission is meso-endemic in Kano State, Nigeria with a prevalence of 32% [22].

### Study setting and population

This study was a cross-sectional study design conducted among selected medical laboratory professionals who were invited for a day of training on malaria microscopy at a primary health care facility, EHA clinics, Kano. A total of 56 Laboratory professionals from both the public and private sectors were involved in this study.

### Training process

In this study, a 1-day training for medical laboratory professionals in Kano was conducted based on the WHO basic microscopy learners guide. The training approach consisted of a series of PowerPoint presentations on implementation issues related to malaria blood film preparation, staining of blood films with Giemsa stain, and examination of high-quality teaching-stained slides for the major Plasmodium species. A pre-test survey was administered to the participants prior to the start of the training to assess their level of knowledge of malaria microscopy; detection, identification, and speciation. Training outcome was measured by the administration of a post-test knowledge assessment survey, and reading a panel of thick-smears to detect malaria parasites.

### Slide preparation and distribution

For this study, already stained slides confirmed to be either malaria-positive or negative were used (5 for each). The slides were prepared and validated by a certified microscopist at EHA Clinics, Kano. Each participant was given 5 slides to view under a light microscope for parasite detection after the training had been completed.

### Questionnaire

A questionnaire including information on the facilities and individuals was electronically sent to invite the participants. The questionnaire was divided into sub-components; the socio-demographic characteristics, educational background, experience, in-service training, routine practice of the professionals, and other facility-related questions. The participants’ knowledge of malaria microscopy before the training was evaluated using a pre-test questionnaire and after the training with a post-test questionnaire.

### Statistical analysis

The Data collected was cleaned and analyzed using R language. Socio-demographic characteristics of the participants and facility-related responses were presented using descriptive statistics. The malaria detection knowledge (median scores) among participants was determined for both the public and private sectors. The significance of malaria detection knowledge improvement in both the private and public sectors was determined using the Wilcoxen Signed-Ranked Test. Sensitivity, specificity, and kappa scores were calculated to assess laboratory professionals’ performance in detecting malaria parasites using light microscopy. The result of the microscopic diagnosis of malaria reported by the participants was evaluated using various parameters. Sensitivity was determined as the ability of participants to diagnose positive blood films; whereas, specificity was calculated for their ability to diagnose negative blood films [23].

### Ethical considerations

The data presented here were collected during a program activity, and formal Research Ethics Committee review was not required. Approval for the study was granted by the Kano State Ministry of Health Research Ethics Committee (approval number: NHREC/17/03/2018). All data were de-identified prior to analysis and the authors had all necessary administrative permissions to access the data.

## Results and discussion

### Socio-demographic of participants

A total of 96 participants responded to the invite survey with 70 (73%) male and 26 (27%) female. 76% of the respondents worked in the public sector while only 24% of them were in the private sector. Most participants had a current practicing license (74%) with more than half of them from the public facilities (54%).

Close to half of the participants held a bachelor’s degree during the time of training (49.5%), 33.6% of them had postgraduate degrees while about one-tenth of them had a diploma (13.7%).

Only a small minority (10.4%) of the participants had attended any malaria training within the past three years. The median year of practice of the participants was 6 years; 4.5 years in the private sector, and 7 years in the public sector.

**Table 1:** Socio-demographic of participants

| **Variables** | **Frequency** |  |  |
| --- | --- | --- | --- |
| **Gender** | **All (n=96)** | Private | Public |
| Male | 70 (73.0%) | 22(46.0%) | 20 (49.0%) |
| Female | 26 (27.0%) | 33 (51.0%) | 21 (51.0%) |
| **Current practicing license** |  |  |  |
| Yes | 71 (74.0%) | 15 (68.2%) | 54 (78.3%) |
| No | 25 (26.0%) | 7 (31.8%) | 15 (21.7%) |
| **Health facility (n=91)** |  | **Private** | **Public** |
|  |  | 22 (24.2%) | 69 (75.8%) |
| **Educational qualification** | **All (n=95)** | **Private** | **Public** |
| Diploma | 13 (13.7%) | 1 (4.3%) | 8 (11.9%) |
| Bachelors | 47 (49.5%) | 15 (65.2%) | 31 (46.3%) |
| Post-graduate | 32 (33.6%) | 7 (30.5%) | 25 (37.3%) |
| Post graduate diploma | 3 (3.2%) | - | 3 (4.5%) |
| **Malaria training within past 3 years** |  |  |  |
| Yes | 10 (10.4%) | 4 (18.2%) | 6 (8.7%) |
| No | 86 (89.6%) | 18 (81.8%) | 63 (91.3%) |
| **Median practice duration (years)** | 6 (17.1%) | 4.5 (12.9) | 7 (20.0%) |

### Facility characteristics of participants

On average, the private facilities conduct about 38 malaria tests per day with a mean positive of 53.4 while the public facilities conduct 81 malaria tests per day with a mean positive of 64.7.

Generally, 70.8% of the participants said that their laboratory was air-conditioned with the majority of them from private facilities (90.3%).

Close to half of the participants used Giemsa stain for slide staining (49.4%), 22.5% of them used field stain A&B while less than one-tenth used Leishman stain. Less than half of them (41.1%) sourced their stain from the scientific store, 24.4% prepared the stains by themselves while 25.6% got them from an open market.

A considerable number of the participants used both RDT and smear for malaria tests in their respective facilities (77.8%), while 13.3% of them used microscopy smear only. The majority that used microscopy smear, air dry slides on the bench (83.5%) while only 16.5% of them used slide dryers.

The plus system was the most adopted method of test result reporting among participants (90%).

**Table 2:** Facility characteristics of participants

| **Variables** | **Frequency** |  |  |
| --- | --- | --- | --- |
| **Number of malaria tests per day (Mean)** | **All (n = 96)** | **Private** | **Public** |
|  | 69.6 | 38.1 | 80.8 |
| **Percentage of positive malaria tests (Mean)** | 62.4 | 53.4 | 64.7 |
| **Lab air-conditioned** |  |  |  |
| Yes | 68 (70.8%) | 20 (90.3%) | 23 (63.8%) |
| No | 28 (29.2%) | 2 (9.1%) | 25 (36.2%) |
| **Type of stain** |  |  |  |
| Giemsa | 44 (49.4%) | 13 (59.1%) | 31 (46.3%) |
| Leishman | 9 (10.1%) | 2 (9.1%) | 7 (10.4%) |
| Field stain A&B | 20 (22.5%) | 7 (31.8%) | 25 (37.3%) |
| Giemsa and Fixed stain A&B | 13 (14.6%) | - | 13 (19.4%) |
| Giemsa and Leishman | 2 (2.3%) | - | 2 (3.0) |
| Leishman and Fixed stain A&B | 1 (1.1%) | - | 1 (1.5%) |
| **Source of stain** |  |  |  |
| Open market | 23 (25.6%) | 4 (18.2%) | 19 (27.9%) |
| Scientific store | 37 (41.1%) | 12 (54.5%) | 25 (36.8%) |
| Self-prepared | 22 (24.4%) | 4 (18.2%) | 18 (26.5%) |
| Open market and self-prepared | 3 (3.3%) | 2 (9.6%) | 1 (1.5%) |
| Others | 6 (6.2%) | - | 5 (7.3%) |
| **Source of RDT** |  |  |  |
| Open market | 11 (13.3%) | - | 8 (12.3%) |
| Scientific store | 18 (21.7%) | - | 12 (18.5%) |
| Supplier | 43 (51.7%) | - | 35 (53.8%) |
| Others | 11 (13.3%) | - | 10 (15.4%) |
| **Type of test** |  |  |  |
| RDT | 8 (8.9%) | 2 (9.1%) | 6 8.8%) |
| Smear | 12 (13.3%) | 5 (22.7%) | 7 (10.3%) |
| Both | 70 (77.8%) | 15 (68.2%) | 55 (80.9%) |
| **Method of drying smear** |  |  |  |
| Air dry on the bench | 76 (83.5%) | 16 (72.7%) | 60 (87.0%) |
| Slide dryer | 15 (16.5%) | 6 (27.3%) | 9 (13.0%) |

| Test result unit |  |  |  |
| --- | --- | --- | --- |
| Plus system | 90 (90.0%) | 20 (90.9%) | 7 (10.3%) |
| Parasites/ul | 9 (10.0%) | 2 (9.1%) | 61 (89.7%) |

### Malaria detection knowledge score among participants

Among participants in the private facilities, the pre-training median knowledge score was 47.5% and the post-training score was 65.0%. Among participants in the public facilities, the pre-training median knowledge score was 55.0% and 70.0% post-training.

The Wilcoxon Signed-Ranked Test was used to determine the level of significance of the improvement in participants’ knowledge after undergoing the training. Participants from the private facilities had an improvement score of 15.0% with V=0 and a p-value of 0.01368. The p-value is less than 0.05 set as the level of significance, this implies that the percentage improvement score of 15.0% seen after the training is statistically significant.

Participants from the public facilities had an improvement score of 20.0% with V=7 and a p-value of 0. The p-value is less than 0.05 set as the level of significance, this implies that the percentage improvement score of 20.0% seen after the training is statistically significant.

**Table 3:** Malaria detection knowledge score among participants

| **Malaria detection knowledge** | **Private** | **Public** | **Both** |
| --- | --- | --- | --- |
| Pre-training median score | 9.5 (47.5%) | 11 (55.0%) | 11 (55.0%) |
| Post-training median score | 13 (65.0%) | 14 (70.0%) | 14 (70.0%) |
| Improvement median score | 4 (20.0%) | 3 (15.0%) | 3 (15.0%) |
| % improvement median score | 15.0% | 20.0% |  |
| Wilcoxon Signed-Rank Test | V = 0, p-value=0.01368 | V = 7, p-value=0 |  |

### Sensitivity, specificity, and agreement of participants in detecting malaria parasite by facility based on the total number of observations

Overall, the sensitivity and specificity of medical laboratory professionals in detecting malaria parasites were 66% and 34% respectively. This current study shows moderate sensitivity but very low specificity which are lower than a study done in Zambia with a sensitivity and specificity of 88% and 91% respectively. This difference could be attributed to the fact that the Zambia study showed the aggregate accuracy in sensitivity and specificity from six healthcare facilities [24]. The findings from our study are also low compared to that from a study in Hawassa, Southern Ethiopia where the sensitivity and specificity were 82% and 96.5% [25]. However, the specificity of malaria detection by laboratory professionals in our study is similar to the 66.3% seen in a study conducted in Haiti [26].

A study conducted by Okoro et al., 2020, in Southeastern Nigeria gave a post-test sensitivity and specificity of 70.4% and 88.3% respectively from a refresher training conducted among laboratory professionals [27]. However, this study had a lower pre-test sensitivity and specificity of 67.4% and 38.9%, which are quite similar to that of our current study.

In our study, the kappa coefficient for agreement on malaria parasite detection between participant readers and an expert reader was found to be 0, indicating a lack of agreement beyond what would be expected by chance alone. This highlights a substantial discrepancy in the detection capabilities between the participants and the expert, pointing to a significant need for enhanced training and a deeper understanding of diagnostic techniques among the participants. This result aligns with observations from a similar study conducted across four laboratories in Mpumalanga province, South Africa, which reported a slight level of disagreement (kappa=0.11), suggesting some challenges in consistency of malaria detection are not uncommon [28]. Conversely, this outcome contrasts with the findings reported by Clendennen et al., where a moderate agreement (kappa=0.61) was observed, indicating variability in diagnostic proficiency and agreement across different studies and settings [29].

Medical laboratory professionals from the private facilities had a sensitivity and specificity of 78% and 33% respectively while professionals from the public facilities had a sensitivity and specificity of 63% and 34% respectively. The professionals from the private facilities had an agreement of kappa=0.12 with the expert reader while professionals from the public facilities had a level of agreement of (kappa= -0.03) with the expert reader. This implies that professionals from the public facility’s level of agreement is worse than what would be expected by a random chance and therefore a high level of disagreement in malaria parasite detections.

The overall Positive Predictive Value (PPV) of 60% indicates that there is a 60% probability that the individuals tested for malaria have the parasite present in their blood. The overall sensitivity of 66% and a PPV of 60% suggest that there is moderate accuracy in malaria parasite detection when the result is positive, however, this is not highly efficient as there is a high significant proportion of false positives and about one-third of the actual positive cases missed. The overall Negative Predictive Value (NPV) of 40% indicates that when the malaria test is negative, there is a 40% chance that the individuals are disease-free. Therefore, the overall specificity of 34% and NPV of 40% indicate a low level of accuracy for ruling out the presence of malaria parasites in the slides examined by the participants. In a study conducted Edo State, Nigeria by Ajakaye & Ibukunoluwa (2020), the sensitivity and specificity of malaria Rapid Diagnostic Tests (RDTs) in comparison to light microscopy were reported as 69.08% and 66.67%, respectively. Furthermore, the study determined the positive predictive value (PPV) and negative predictive value (NPV) to be 99.6% and 1.77%, respectively [30].

**Table 4:**
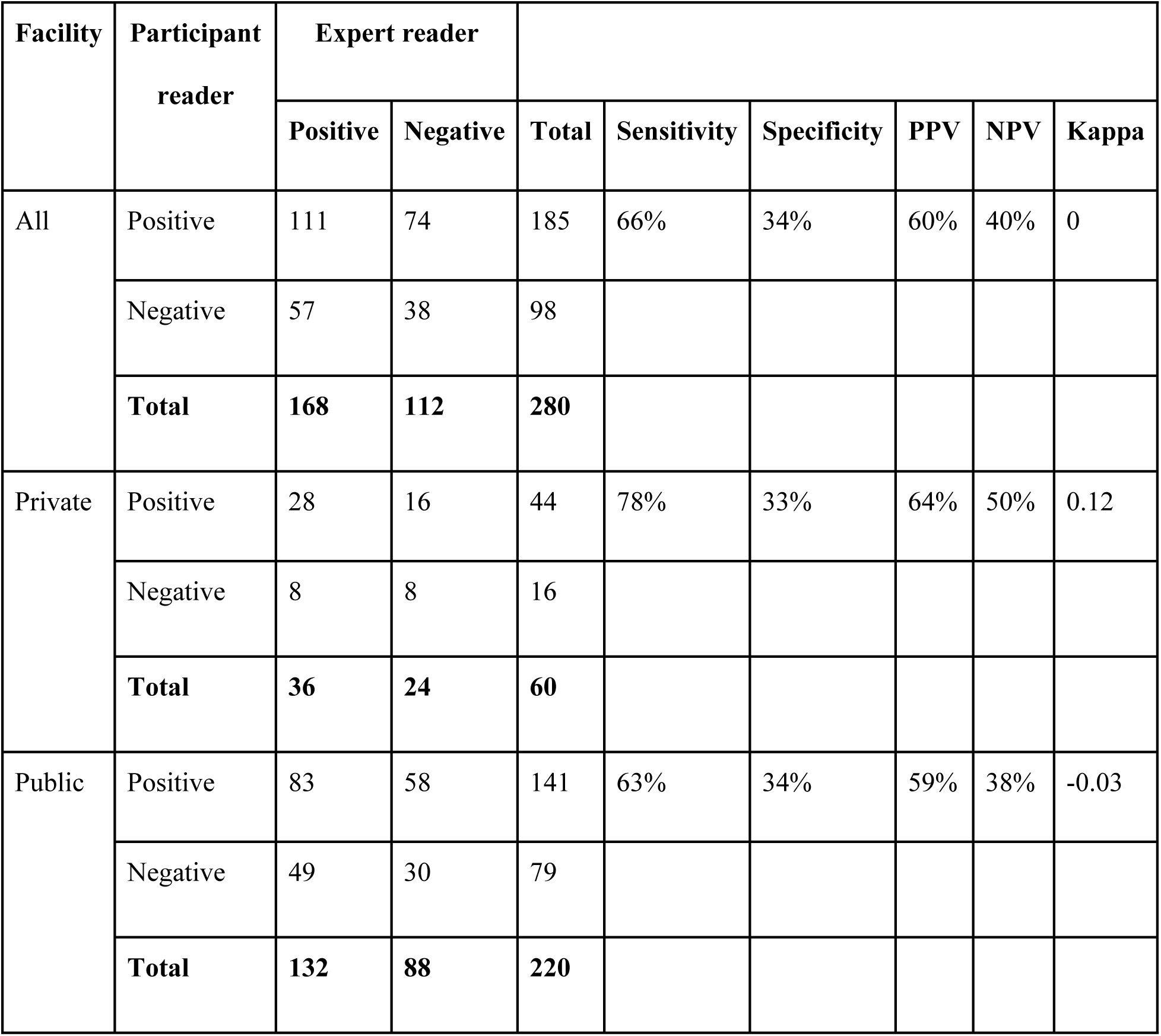
Sensitivity, specificity, and agreement of participants in detecting malaria parasite by facility based on the total number of observations.

## Limitations

The limitation of this study is that rather than assessing routine or daily performance in the diagnosis of malaria, we only employed proficiency testing slides to assess laboratory experts’ expertise under ideal circumstances. Additionally, we did not assess the laboratory staff’s competence in terms of smear preparation and blood film staining for the diagnosis of malaria. The sample size of 56 is also considered a limitation as this may not truly represent laboratory professionals in Kano Metropolis.

## Conclusion

There was a significant improvement in the malaria detection knowledge based on the improvement score from pre-training and post-training assessments. However, there was low sensitivity and specificity in the detection of malaria parasites as well as a low level of agreement of the participants with expert microscopists in the detection of malaria parasites. There is poor agreement in the detection of malaria parasites in participants from the public facilities compared to the participants from the private facilities. Microscopy is the golden standard for malaria diagnosis, but it requires an expert microscopist to obtain high levels of sensitivity, so it is essential to invest in regular training and the qualification of these professionals.

## Authors’ contributions

● Nirmal Ravi: The contribution of this author to the manuscript involved initial conception of research goals and aims, significant oversight and leadership throughout the planning and execution phases of the research activity and played a critical role in the preparation and refinement of this manuscript.
● Oluwaseunayo Deborah Ayando: The author significantly contributed to the manuscript through the application of rigorous formal techniques, including statistical methodology, to analyze and synthesize the study data effectively. This analytical process was crucial in interpreting the research findings accurately and laying the groundwork for the study’s conclusions. Furthermore, the author was pivotal in the creation and development of the published work, taking the lead in writing the initial draft. This task involved not only the articulation of the research findings but also the substantive translation of complex data into accessible and coherent narratives, ensuring the study’s contributions were clearly communicated and understood.
● Okoronkwo Marizu Obinnaya : The author took on a significant management and coordination role, overseeing the research activity’s planning and execution. This included organizing and conducting training sessions, which were crucial for ensuring the research team’s proficiency and effectiveness. Through these efforts, the author ensured that the research was conducted efficiently, adhering to the highest standards of quality and integrity.

## Data Availability

All relevant data are within the manuscript and its Supporting Information files.

## Acknowledgement

We extend our sincere gratitude to the management of EHA Clinics and the Laboratory unit for their invaluable assistance and support, which played a pivotal role in the execution of our study. Special appreciation goes to the laboratory professionals who participated in our research; their expertise and commitment were fundamental to the integrity and success of our study.

